# Geographical variation and factors associated with unsafe child stool disposal in Ethiopia: A spatial and multilevel analysis

**DOI:** 10.1101/2020.09.07.20189902

**Authors:** Biniyam Sahiledengle, Zinash Teferu, Yohannes Tekalegn, Tadesse Awoke, Demisu Zenbaba, Kebebe Bekele, Abdi Tesemma, Fikadu Seyoum, Demelash Woldeyohannes

## Abstract

**Background:** Unsafe disposal of children’s stool makes children susceptible to fecal-oral diseases and children remain vulnerable till the stools of all children are disposed of safely. There is a paucity of data on spatial distribution and factors associated with unsafe child stool disposal in Ethiopia. Previous estimates, however, do not include information regarding individual and community-level factors associated with unsafe child stool disposal. Hence, the current study aimed (i) to explore the spatial distribution and (ii) to identify factors associated with unsafe child stool disposal in Ethiopia.

**Methods:** A secondary data analysis was conducted using the recent 2016 Ethiopian demographic and health survey data. A total of 4145 children aged 0–23 months with their mother were included in this analysis. The Getis-Ord spatial statistical tool was used to identify high and low hotspots areas of unsafe child stool disposal. The Bernoulli model was applied using Kilduff SaTScan version 9.6 software to identify significant spatial clusters. A multilevel binary logistic regression model was fitted to identify factors associated with unsafe child stool disposal.

**Results:** Unsafe child stool disposal was spatially clustered in Ethiopia (Moran’s Index = 0.211, p-value < 0.0001), and significant spatial SaTScan clusters of areas with a high rate of unsafe child stool disposal were detected. The most likely primary SaTScan cluster was detected in Tigray, Amhara, Afar (north), and Benishangul-Gumuz (north) regions (LLR: 41.62, p< 0.0001). Unsafe child stool disposal is more prevalent among households that had unimproved toilet facility (AOR = 1.54, 95%CI: 1.17-2.02), and those with high community poorer level (AOR: 1.74, 95%CI: 1.23-2.46). Higher prevalence of unsafe child stool disposal was also found in households with poor wealth quintiles. Children belong to agrarian regions (AOR: 0.62, 95%CI 0.42-0.91), children 6-11 months of age (AOR: 0.66, 95%CI: 0.52-0.83), 12-17 months of age (AOR: 0.68, 95%CI: 0.54-0.86), and 18-23 months of age (AOR: 0.58, 95%CI: 0.45-0.74) had lower odds of unsafe child stool disposal.

**Conclusions:** Unsafe child stool disposal was spatially clustered. Higher odds of unsafe child stool disposal were found in households with high community poverty level, poor, unimproved toilet facility, and with the youngest children. Hence, the health authorities could tailor effective child stool management programs to mitigate the inequalities identified in this study. It is also better to consider child stool management intervention in existing sanitation activities considering the identified factors.

## Background

Disposal of child stool in the open field, thrown into the garbage, put into open drains, burying, or left on the ground are considered unsafe [1,2]. Unsafe disposal of children’s stool makes children susceptible to many fecal-oral diseases [1,3–5]. A systematic review by Gil et al. reported that unsafe child stool disposal associated with a 23% (RR: 1.23, 95% CI: 1.15-1.32) increase in the risk of diarrheal diseases in children [6]. Another review of case-control studies showed that the disposal of a child’s feces into a latrine decreases the odds of diarrhea by about 25% in children under five years of age (OR: 0.73, 95% CI: 0.62-0.85) [7]. Furthermore, studies reported the effect of young children’s stool disposal and increased risk of stunting [8,9].

In Ethiopia, unsafe child stool disposal is a huge challenge and Ethiopia ranked number 26 for the percentage of children whose feces are safely disposed of, putting the country among the worst third of 38 African countries with available Multiple Indicator Cluster Survey (MICS) [2]. And diarrhea is one of the major contributors to deaths for under age 5 children in the country [10]. Based on the WHO/CHERG estimates, diarrhea contributes to more than one in every ten (13%) child deaths in Ethiopia [11]. According to a recent pooled data analysis using DHS surveys in Ethiopia, 77 percent of children feces disposed of unsafely; either throw outside the yard or not disposed of [12].

Previous studies identified multiple factors that contribute to the occurrence of unsafe child stool disposal [12–20]. Socio-demographic (age of the child, sex of the child, age of the mother, maternal educational status, place of residence) [12–14,21], socio-economic (household wealth index) [14,17], and access to sanitation facility [12–14,17,19] were associated with unsafe child stool disposal practices. Moreover, given the fact that unsafe child stool disposal is associated with open defecation, the prevalence of unsafe child stool disposal practices in rural areas is higher compared to urban areas [12,22]. In the context of this research, higher rates of unsafe child stool disposal were found in poor, rural households with the youngest children and where other household members defecate in the open [14–22].

So far few studies were conducted on child stool disposal in Ethiopia [12–14], and previous estimates, however, identified the determinants of child stool disposal using a standard logistic regression model that does not include information regarding individual and community-level factors [12–14]. Thus, a multilevel regression model is required, which considers the hierarchal and cluster nature of the Ethiopian Demographic and Health Survey (EDHS) data and enhances the accuracy of estimates. And to date, studies on child stool disposal in Ethiopia have not assessed the spatial distribution of unsafe child stool disposal. Therefore, the current study aimed (i) to explore the spatial distribution and (ii) to identify factors associated with unsafe child stool disposal in Ethiopia using a multilevel regression model.

## Methods

### Data source and sampling

A secondary data analysis of the Ethiopia Demographic and Health Survey (EDHS) conducted in the year 2016 was used. The EDHS-2016 is the recent survey implemented by the Central Statistical Agency (CSA). The survey was conducted from January 18, 2016, to June 27, 2016, based on a nationally representative sample that provides estimates at the national and regional levels and for urban and rural areas.

The surveys used a stratified two-stage cluster sampling technique. The sampling frame used for the EDHS-2016 is the Ethiopia Population and Housing Census (PHC), which was conducted in 2007 by the Ethiopia Central Statistical Agency. The census frame is a complete list of 84,915 enumeration areas (EAs) created for the 2007 PHC. An EA is a geographic area covering on average 181 households. In the first stage, a total of 645 EAs (202 in urban areas and 443 in rural areas) were selected with probability proportional to EA size and with independent selection in each sampling stratum. In the second stage of selection, a fixed number of 28 households per cluster were selected with an equal probability systematic selection from the newly created household listing [10]. The present study included the youngest child under age 2 living with the mother.

### Data source and extraction

The EDHS-2016 data sets were downloaded in STATA format with permission from the Measure DHS website (http://www.dhsprogram.com). Unsafe child stool disposal and its potential predictor variables at the individual and community levels were extracted accordingly.

### Study variables

#### Outcome variable

The outcome variable for this study was child stool disposal, which is dichotomized as unsafe and safe. A child’s stool was considered to be disposed of “safely” when the child used latrine/ toilet or child’s stool was put/rinsed into a toilet/latrine, whereas other methods were considered “unsafe” (i.e. put, rinsed in a drain, ditch, thrown in the garbage, left or buried in the open). The survey collected these data from mothers’ verbal reports on whether the child’s last stools were put in or rinsed into a toilet or latrine, buried, or the child used a toilet or latrine.

#### Independent variables

The independent variables for this study were classified as individual and community level factors. The individual-level variables of this study were (age of the child, sex of the child, presence of diarrhea in the last two weeks, the source of drinking water, sanitation facilities, mother educational level, mother occupation, and household wealth quintile). The contextual region, place of residence, and community poverty were identified as community-level variables (**Table 1**). The choice of independent variables was guided by the previous works of literature [12–15]. Community poverty level (proportion of women in the poorest and poorer quintile derived from data on wealth index which is categorized as low and high poverty community). The interest of the current study was not in the regions delineated for administrative purposes, which might not necessarily be related to child stool disposal of the population. Accordingly, in the current study, the regions were categorized into agrarian, pastoralist, and city. The regions of Tigray, Amhara, Oromiya, SNNP, Gambella, and Benshangul Gumuz were recorded as agrarian. The Somali and Afar regions were combined to form the pastoralist region and the city administrations-Addis Ababa, Dire Dawa, and Harar were combined as the city.

**Table 1:** Independent variables and categorization

| Individual-level factors (Level 1) | Category |
| --- | --- |
| **Child characteristics** |  |
| Sex of the child | Categorized in to (1) male; or (2) female |
| Age of child (months) | Categorized into (1) 0–5; (2) 6–11; (3) 12–17; and (4) 18–23 |
| Diarrhea in the last two weeks | Categorized into (1) yes; or (2) no |
| **Maternal/paternal/household Characteristics** |  |
| Maternal age in years | Categorized into (1) 15-24; (2) 25-34; and $\geq 35$ |
| Educational level of mother | Categorized in to (1) no formal education; (2) primary; (3); secondary; or (4) higher |
| Mother's working status | Categorized in to (1) not working, or (2) working |
| Number of under-five children | Categorized into (1) $\leq 2$; or (2) $\geq 3$ |
| Wealth index | Categorized into (1) (first quintile) (poorest); (2) (second quintile) (poorer); (3) (third quintile) (middle); (4) (fourth quintile) (richer); or (5) (fifth quintile) (richest) |
| Source of drinking water | Categorized in to (1) improved; or (2) unimproved |
| Latrine type | Categorized in to (1) improved; or (2) unimproved |
| **Community-level factors (Level 2)** |  |
| Place of residence | Categorized into (1) urban; or (2) rural |
| Region | Categorized in to (1) agrarian; (2) pastoralist or (3) city |
| Community poverty | Categorized in to (1) high; or (2) low |

### Data management and analysis

Descriptive measures were used to illustrate the overall characteristics of the study participants. A sampling weight was done to adjust for the non-proportional allocation of the sample to different regions and the possible differences in response rates. A detailed explanation of the weighting procedure can be found in the EDHS methodology parts [10].

In Ethiopian DHS data, children within a cluster more similar to each other than between clusters. When analyzing such datasets, a multilevel model is generally more appropriate than the standard regression model because it enables one to deal with the hierarchical structure of variables. For this reason, a multilevel model was used to identify factors associated with unsafe child stool disposal. As the response variable was dichotomous (safe, unsafe), multilevel binary logistic regression was fitted. The model goodness of fit was checked using deviance and Akakie Information Criteria (AIC). The model with the lowest deviance and AIC was chosen. The Proportional Change in Variance (PCV) was computed for each model with respect to the empty model to show the power of the factors in the model to explain unsafe child stool disposal. Accordingly, the PCV was calculated by the following formula [PCV = (Ve-Vmi)/Ve], where Ve is variance in unsafe child stool disposal in the empty model and Vmi is variance in successive models. [Median Odds Ratio 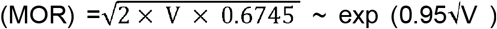], where V is the estimated variance of clusters. The MOR measure is always greater than or equal to 1. If the MOR is 1, there is no variation between clusters. Variables with p-value < 0.25 in the bi-variable analysis were fitted in the multivariable model. Adjusted Odds Ratio (AOR) with a 95% Confidence Interval (CI) and p-value < 0.05 in the multivariable model were used to declare significant association with unsafe child stool disposal. Variance inflator factor (VIF) was employed for checking multicollinearity among the independent variables. STATA version 14 software was used for all statistical analyses.

### Spatial autocorrelation analysis

In this study, the spatial analysis was performed using the spatial statistics tool (ArcGIS Version 10.3; Redlands, California, United States). The spatial autocorrelation (Global Moran’s I) statistic measures were used to evaluate whether unsafe child stool disposal was dispersed, clustered, or randomly distributed [23]. Spatial heterogeneity of high /low areas of unsafe child stool disposal was examined using the Getis-Ord Gi* statistics and associated Z-scores. Moreover, the spatial interpolation technique was applied (using the ordinary kriging interpolation technique) to predict the unsampled /unmeasured value from sampled measurements.

### Spatial scan statistical analysis

Spatial scan statistical analysis was employed to identify the geographical locations of statistically significant spatial clusters of unsafe child stool disposal in Ethiopia using SaTScan™ version 9.6 software. Unsafe child stool was taken as cases and those with safe child stool disposal as controls to fit the Bernoulli model [24]. The default maximum spatial cluster size of < 50% of the population was used. A Likelihood ratio test statistic was used to determine whether the number of observed unsafe stool disposal cases within the potential cluster was significantly higher than the expected or not. Primary and secondary clusters were identified using p-values and log-likelihood ratio tests.

## Results

### Socio-demographic characteristics of participants

A total of 4145 children aged 0-23 months with their mother were included in this analysis. Of these, 2164 (52.2%) were female with a mean age of 10.66 months (SD ± 0.11). The majority of 3647 (88.0%) of the respondents were rural residents. About 2500 (60.3%) of the children’s mother had no formal education and about one-fifth were in the poorest wealth quintile (**Table 2**).

**Table 2:** Socio-demographic and socio-economic characteristics of study participants, EDHS 2016 (N = 4145)

| Background characteristics | Weighted frequency (n) | % |
| --- | --- | --- |
| **Individual-level factors** |  |  |
| **Sex of the child (n=4144)** |  |  |
| Male | 1980 | 47.8 |
| Female | 2164 | 52.2 |
| **Age of the child** |  |  |
| 0-5 months | 1059 | 25.6 |
| 6-11 months | 1085 | 26.2 |
| 12-17 months | 814 | 19.6 |
| 18-23 months | 1187 | 28.6 |
| **Diarrhea in the last two weeks (n=4129)** |  |  |
| Yes | 670 | 16.2 |
| No | 3459 | 83.8 |
| **Mother educational level** |  |  |
| No formal education | 2500 | 60.3 |
| Primary | 1279 | 30.9 |
| Secondary | 254 | 6.1 |
| Higher | 112 | 2.7 |
| **Mother's age (4143)** |  |  |
| 15-24 | 1215 | 29.3 |
| 25-34 | 2105 | 50.8 |
| >34 | 823 | 19.9 |
| **Mother's working status** |  |  |
| Not working | 2439 | 58.85 |
| Working | 1705 | 41.15 |
| **Number of under-five children** |  |  |
| 1-2 | 3458 | 83.43 |
| $\geq 3$ | 687 | 16.57 |
| **Household wealth index** |  |  |
| Poorest | 975 | 23.5 |
| Poorer | 905 | 21.8 |
| Middle | 867 | 20.9 |
| Richer | 754 | 18.2 |
| Richest | 642 | 15.5 |
| **Toilet facility** |  |  |
| Improved <sup>a</sup> | 419 | 10.1 |
| Unimproved | 3726 | 89.9 |
| **Source of drinking water** |  |  |
| Improved <sup>b</sup> | 2330 | 56.2 |
| Unimproved | 1815 | 43.8 |
| **Community-level factors** |  |  |
| **Region** |  |  |
| Agrarian | 3802 | 91.74 |
| Pastoralist | 210 | 3.19 |
| City | 132 | 5.07 |
| **Place of residence** |  |  |
| Urban | 498 | 12.0 |
| Rural | 3647 | 88.0 |
| **Community poverty level** |  |  |
| High | 1647 | 39.74 |
| Low | 2498 | 60.26 |

<sup>a</sup>Facilities that would be considered improved if they were not shared by two or more households
<sup>b</sup> Include piped water, public taps, standpipes, tube wells, boreholes, protected dug wells and springs, rainwater, and bottled water.

### Unsafe child stool disposal status

The prevalence of unsafe child stool disposal was 63.10% (95%CI: 59.5-66.6%). Unsafe child stool disposal is varied across urban-rural areas, age of the child, and household wealth index (**Table 3**).

**Table 3:** Binary multilevel logistic regression analysis to determine associated factors of unsafe child stool disposal in Ethiopia, EDHS 2016

| Background characteristics | Child stool disposal |  | Crude OR (95%CI) | p-value |
| --- | --- | --- | --- | --- |
|  | Unsafe | Safe |  |  |
| **Individual-level factors** |  |  |  |  |
| **Sex of the child (n=4144)** |  |  |  |  |
| Male | 1226 | 754 | 1 |  |
| Female | 1272 | 892 | 0.84(0.72-0.98) | 0.037 |
| **Age of the child** |  |  |  |  |
| 0-5 months | 401 | 786 | 1 |  |
| 6-11 months | 456 | 604 | 0.63 (0.51-0.79) | p<0.001 |
| 12-17 months | 444 | 641 | 0.69(0.55-0.86) | 0.001 |
| 18-23 months | 346 | 467 | 0.54(0.42-0.69) | p<0.001 |
| **Diarrhea in the last two weeks (n=4129)** |  |  |  |  |
| Yes | 352 | 318 | 0.76(0.61-0.95) | 0.017 |
| No | 2133 | 1326 | 1 |  |
| **Mother educational level** |  |  |  |  |
| No formal education | 1618 | 882 | 1 |  |
| Primary | 715 | 564 | 0.66(0.54-0.80) | p<0.001 |
| Secondary | 128 | 126 | 0.69(0.50-0.94) | 0.018 |
| Higher | 38 | 74 | 0.42(0.27-0.64) | p<0.001 |
| **Mother's age (4143)** |  |  |  |  |
| 15-24 | 793 | 422 | 1 |  |
| 25-34 | 1227 | 878 | 0.85(0.70-1.03) | 0.093 |
| >34 | 477 | 346 | 0.87(0.68-1.10) | 0.248 |
| **Mother's working status** |  |  |  |  |
| Not working | 1577 | 862 | 1 |  |
| Working | 1038 | 668 | 0.81(0.67-0.97) | 0.020 |
| **Number of under-five children** |  |  |  |  |
| 1-2 | 2167 | 1291 | 1 |  |
| ≥ 3 | 448 | 239 | 1.07(0.85-1.33) | 0.558 |
| **Household wealth index** |  |  |  |  |
| Poorest | 771 | 204 | 8.02(6.01-10.73) | p<0.001 |
| Poorer | 598 | 307 | 3.85(2.84-5.22) | p<0.001 |
| Middle | 494 | 373 | 2.60(1.92-3.54) | p<0.001 |
| Richer | 391 | 363 | 1.84(1.35-2.49) | p<0.001 |
| Richest | 243 | 399 | 1 |  |
| **Toilet facility** |  |  |  |  |
| Improved* | 198 | 221 | 1 |  |
| Unimproved | 2300 | 1426 | 2.53(1.99-3.21) | p<0.001 |
| **Source of drinking water** |  |  |  |  |
| Improved** | 1298 | 1032 | 1 |  |
| Unimproved | 1200 | 615 | 1.76(1.42-2.16) | p<0.001 |
| **Community-level factors** |  |  |  |  |
| **Region** |  |  |  |  |
| Agrarian | 2387 | 1415 | 1.65(1.16-2.38) | 0.006 |
| Pastoralist | 159 | 51 | 4.61(2.91-7.30) | p<0.001 |
| City | 69 | 63 | 1 |  |
| **Place of residence** |  |  |  |  |
| Urban | 193 | 305 | 1 |  |
| Rural | 2305 | 1342 | 3.89(2.85-5.30) | p<0.001 |
| **Community poverty level** |  |  |  |  |
| High | 1374 | 1124 | 5.05(3.84-6.62) | p<0.001 |
| Low | 1241 | 406 | 1 |  |

### Spatial distribution of unsafe child stool disposal in Ethiopia

The analysis of spatial autocorrelation indicated that the spatial distribution of unsafe child stool disposal was clustered in Ethiopia. The Global Moran’s I value 0.211 (p-value < 0.0001) indicated that there was significant clustering of unsafe child stool disposal in Ethiopia (**Additional File 1**).

Hot-spot areas were found in Tigray (Central, and northeast), Amhara (Central, North, and Southeast), Afar (West, and South), Gambela (West), Oromia (South and East), North and Southeast parts of Somali regions, while cold-spot areas were found in SNNP (North, West, and East), Benishangul-Gumuz (Southwest), Addis Ababa, Harari and Dire Dawa (**Figure 1**).

**Figure 1:**
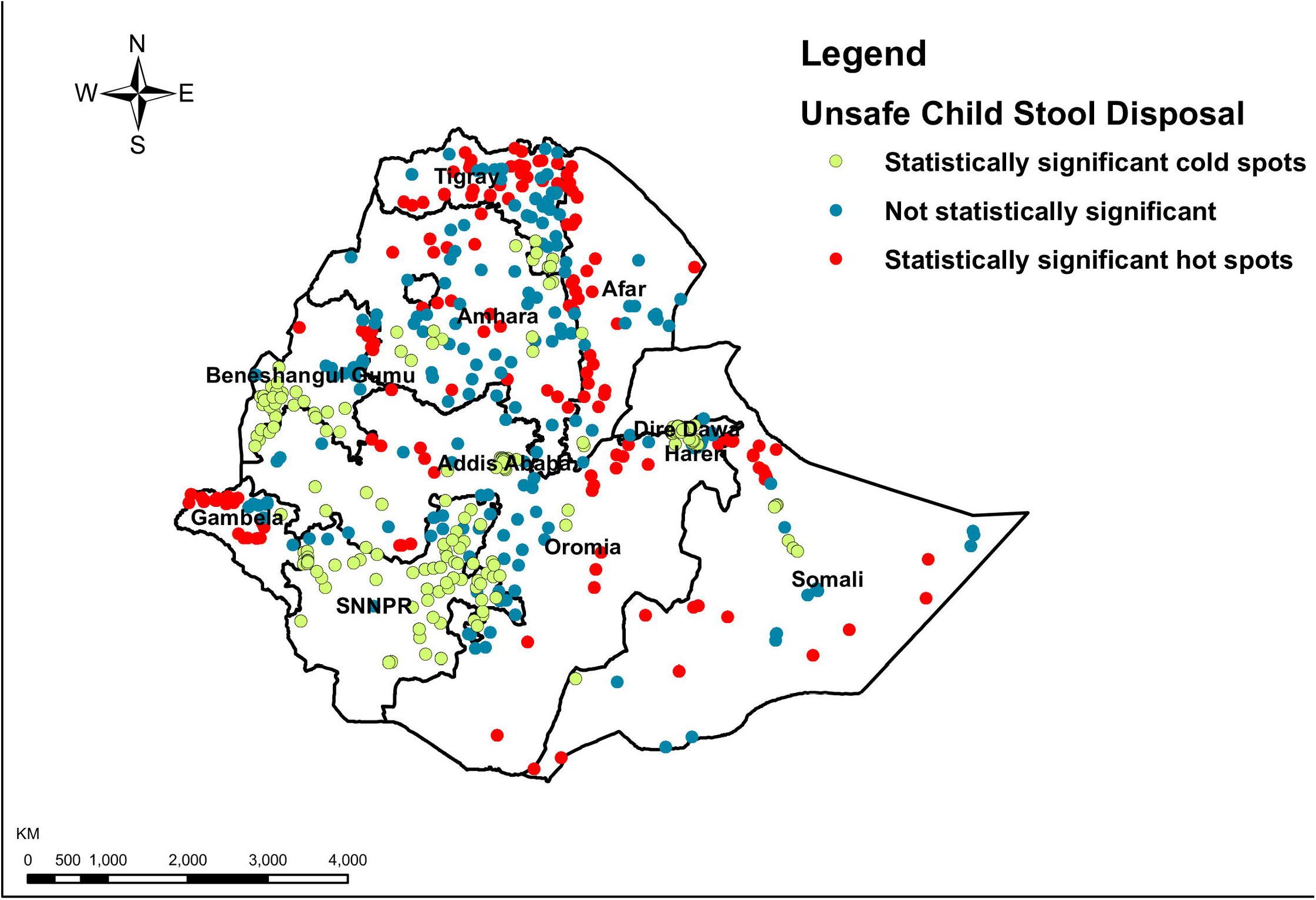
Hotspot and cold spot analysis using Getis-Ord Gi statistics of unsafe child stool disposal in Ethiopia: A single dot on the map represents one enumeration area, EDHS 2016.

Ordinary kriging interpolation analysis was conducted to predict child stool disposal in Ethiopia. High unsafe child stool disposal areas were found in Tigray, Amhara, Afar, Gambela, Southern Somali, and Southeastern parts of Oromia regions. In contrast, low unsafe child stool disposal areas were predicted in SNNP, Southern parts of Benishangul-Gumuz, Northern Somali, Western Oromia, and some parts of Amhara regions (**Figure 2**).

**Figure 2:**
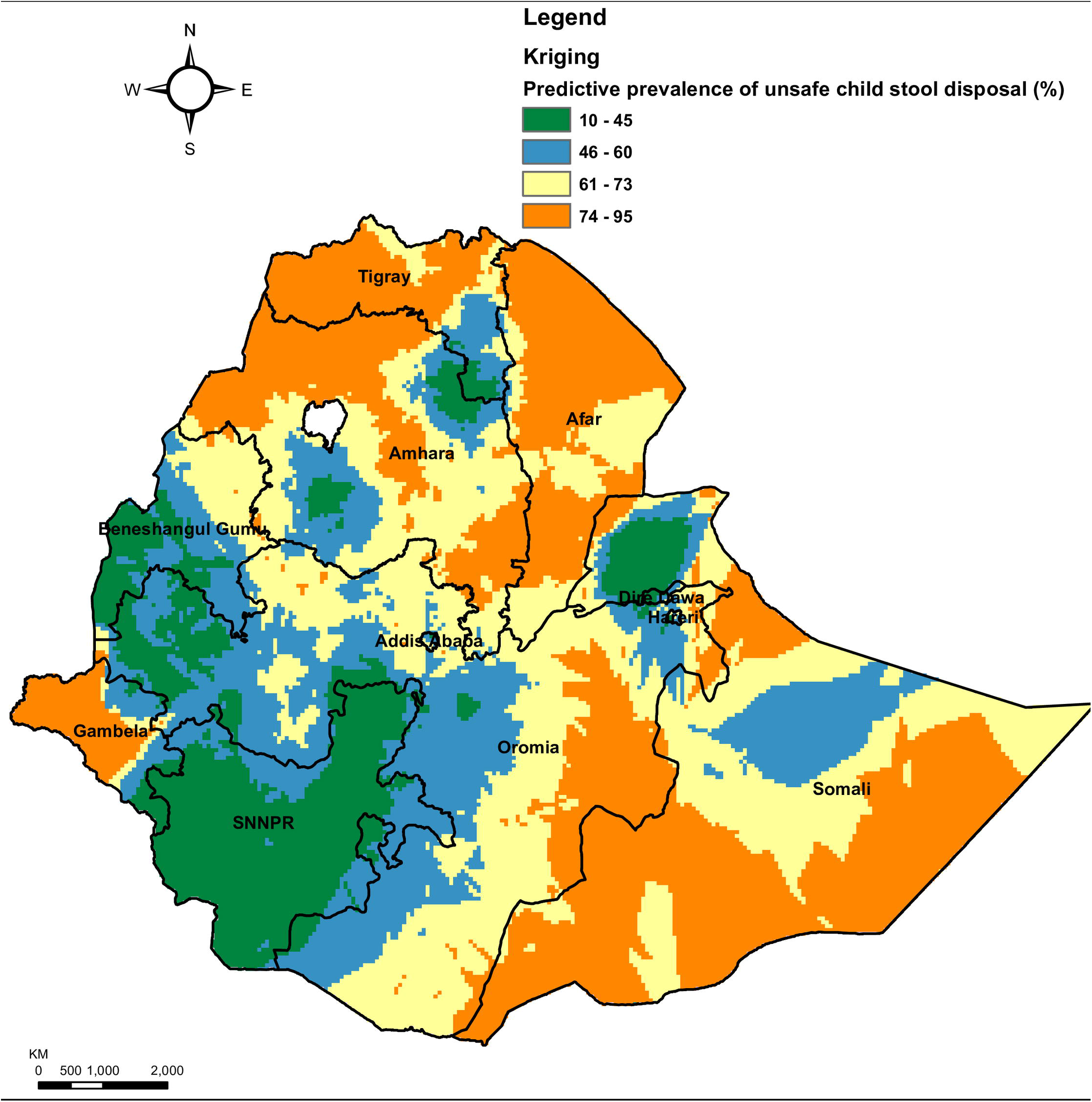
Ordinary kriging interpolation of unsafe child stool disposal in Ethiopia, EDHS 2016.

### Spatial scan statistical analysis

**Table 4** show significant spatial clusters of unsafe child stool disposal in Ethiopia. Most likely (primary clusters) and secondary clusters of unsafe child stool disposal were identified. A total of 270 significant clusters were identified at which 201 were most likely (primary) and 69 secondary clusters. The primary clusters were located in Tigray, Amhara, and Afar regions. The primary clusters were centered at 13.351814 N, 38.353591 E with 471.07 km radius, a relative risk (RR) of 1.26, and the Log-Likelihood Ratio (LLR) of 41.62, at p< 0.0001. The bright pink colors indicate that the most statistically significant spatial windows contain primary clusters of unsafe child stool disposal in Ethiopia. There was high unsafe child stool disposal within the cluster than outside the cluster (**Figure 3**).

**Figure 3:**
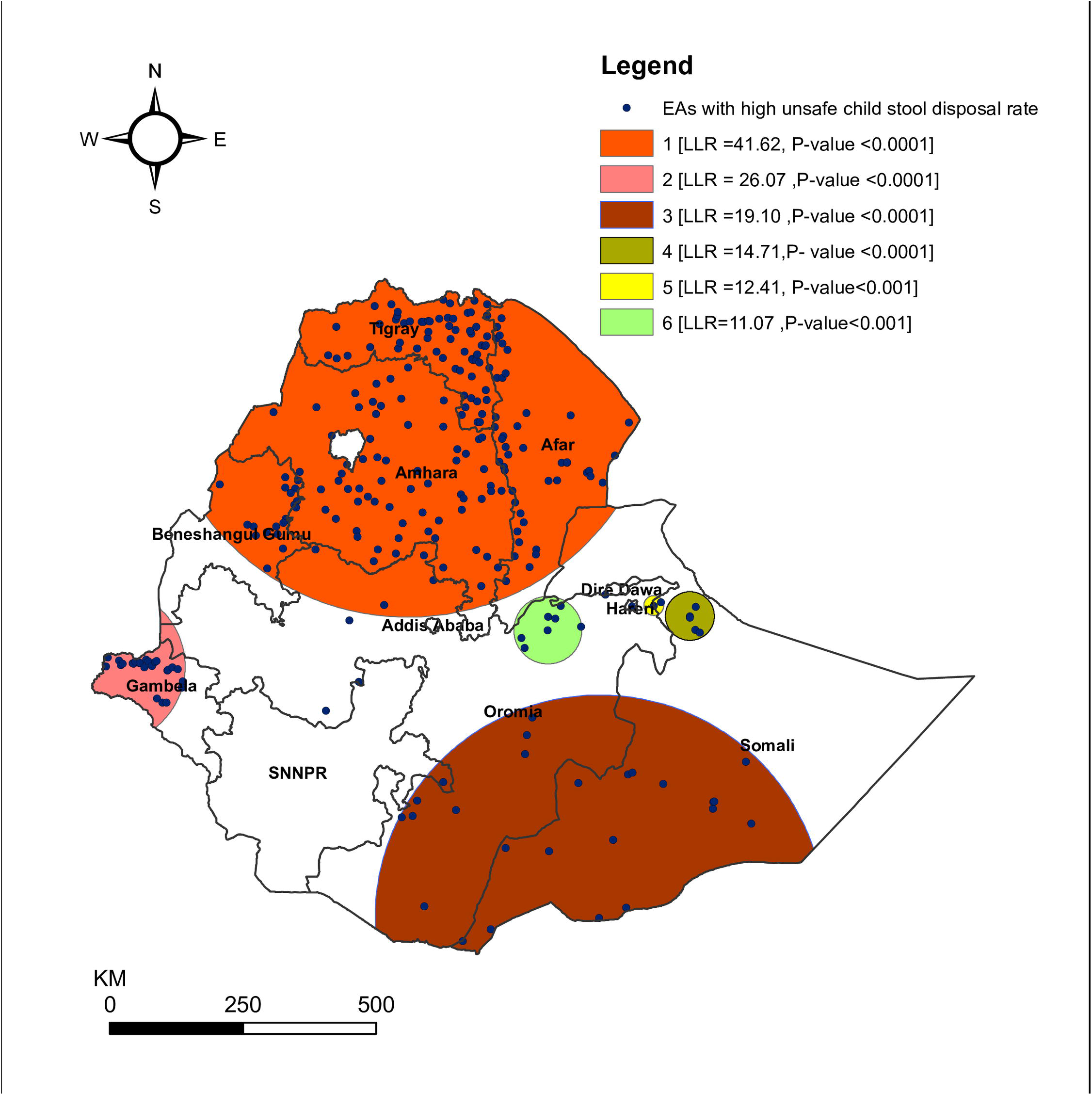
The spatial clustering of areas with high unsafe child stool disposal in Ethiopia, EDHS 2016.

**Table 4:** Significant spatial clusters of unsafe child stool disposal in Ethiopia, EDHS 2016

| Clusters (n) | Enumeration areas(clusters) detected | Coordinates/radius | Population | Cases | RR | LLR | P-value |
| --- | --- | --- | --- | --- | --- | --- | --- |
| 1 (201) | 425, 80, 551, 188, 340, 628, 579, 156, 322, 575, 636, 258, 181, 584, 152, 538, 597, 400, 542, 312, 98, 590, 81, 355, 424, 583, 255, 327, 528, 199, 84, 430, 481, 638, 604, 612, 392, 640, 45, 160, 237, 94, 550, 78, 461, 605, 66, 136, 268, 220, 143, 384, 296, 300, 129, 226, 479, 623, 89, 449, 341, 99, 298, 504, 79, 598, 163, 421, 253, 404, 196, 128, 442, 97, 511, 351, 127, 413, 117, 192, 130, 512, 401, 132, 362, 235, 172, 263, 279, 585, 488, 200, 103, 455, 292, 158, 478, 627, 591, 249, 134, 169, 456, 344, 332, 73, 545, 38, 544, 431, 241, 599, 496, 167, 189, 389, 516, 403, 382, 354, 571, 24, 348, 120, 191, 52, 616, 429, 611, 176, 345, 361, 259, 18, 617, 254, 206, 460, 602, 368, 415, 109, 541, 10, 3, 427, 482, 386, 375, 548, 55, 267, 515, 474, 615, 498, 205, 570, 547, 531, 499, 178, 334, 350, 510, 276, 533, 310, 246, 218, 620, 559, 572, 494, 637, 36, 440, 283, 632, 150, 596, 423, 183, 102, 295, 184, 256, 137, 37, 4, 35, 75, 244, 135, 366, 201, 336, 484, 517, 320, 399 | (13.351814 N, 38.353591 E) / 471.07 km | 1,272 | 931 | 1.26 | 41.62 | < 0.0001 |
| 2<sup>a</sup> (26) | 618, 266, 309, 435, 536, 370, 507, 592, 260, 104, 233, 69, 426, 603, 346, 315, 13, 567, 343, 105, 417, 284, 106, 265, 593, 270 | (8.238420 N, 33.229506 E) / 147.69 km | 164 | 144 | 1.41 | 26.07 | < 0.0001 |
| 3<sup>b</sup> (26) | 208, 520, 556, 394, 7, 377, 480, 82, 187, 278, 318, 164, 358, 85, 601, 138, 422, 289, 472, 34, 452, 398, 492, 316, 21, 146 | (4.006703 N, 41.599741 E) / 419.89 km | 256 | 206 | 1.30 | 19.10 | <0.0001 |
| 4<sup>c</sup> (6) | 277, 568, 527, 22, 116, 33 | (9.107168 N, 43.165843 E) / 45.70 km | 41 | 40 | 1.55 | 14.71 | <0.0001 |
| 5<sup>d</sup> (3) | 186, 8, 210 | (9.292185 N, 42.553365 E) / 18.63 km | 27 | 27 | 1.59 | 12.41 | <0.001 |
| 6<sup>e</sup> (8) | 476, 506, 412, 122, 333, 245, 372, 529 | (8.888553 N, 40.744565 E) / 63.62 km | 81 | 70 | 1.38 | 11.07 | <0.001 |

<sup>a</sup>The secondary clusters' were typically located in the Gambela region and centered at 8.238420 N, 33.229506 E with 147.69 km radius, RR: 1.41, and LLR of 26.07 at $p$ -value < 0.0001. It shows that children within the area had 1.41 times higher risk of unsafe child stool disposal than outside the area.
<sup>b</sup>The third clusters' were located in Oromia (south) and Somali (southeast) regions and centered at 4.006703 N, 41.599741 E with 419.89 km radius, RR: 1.30 and LLR of 19.10 at $p$ -value < 0.0001. <sup>c</sup>The fourth clusters' were located in Somali (north) regions and centered at 9.107168 N, 43.165843 E with 45.70 km radius, RR: 1.55, and LLR of 14.41 at $p$ -value < 0.0001. <sup>d</sup>The fifth clusters' were typically located in Hareri regions and centered at 9.292185 N, 42.553365 E with 18.63 km radius, RR: 1.59, and LLR of 12.41 at $p$ -value < 0.001. <sup>e</sup>The six clusters' were typically located in Oromia (northeast) regions and centered at 8.888553 N, 40.744565 E with 63.62 km radius, RR: 1.38, and LLR of 11.07 at $p$ -value < 0.001.

### Measures of variation (random-effects) and model fit statistics

**Table 5** shows the measures of variation (random intercept models) and model fit statistics. Model comparison was done using deviance. The comparison was done among model with no independent variables (the null model), model 1 (a model with only individual-level factors), model 2 (a model with only community-level factors), and model 3 (a model with both individual and community level independent variables simultaneously). A model with the lowest deviance (model 3) was selected. According to the multilevel logistic regression model, about 39.61 % of the variance in the odds of unsafe child stool disposal could be attributed to community-level factors. And the between-cluster variability declined over successive models, from 39.61% in the empty model to 29.62% in the combined model. In the final model (model 3), individual and community-level factors accounted for about 35.87% of the variation observed for unsafe child stool disposal.

**Table 5:** Measures of variation (random intercept models) and model fit statistics for unsafe child stool disposal in Ethiopia, EDHS 2016

| Individual- and community-level characteristics | Null model<br>Empty mode | Model 1<sup>b</sup><br>Individual-level variables<br>AOR (95% CI) | Model 2<sup>c</sup><br>Community-level variables<br>AOR (95% CI) | Model 3<sup>d</sup><br>Individual- and community-level variables<br>AOR (95% CI) |
| --- | --- | --- | --- | --- |
| **Random effect** |  |  |  |  |
| Community-level variance (SE) | 2.155(0.082)*** | 1.394(0.075)*** | 1.442(0.074)*** | 1.382(0.074)*** |
| ICC (%) | 39.61% | 29.78% | 30.51% | 29.62% |
| MOR<sup>e</sup> | 4.03 | 3.06 | 3.12 | 3.05 |
| PCV (%) | Reference | 35.31 | 33.08 | 35.87 |
| **Model fit statistics** |  |  |  |  |
| AIC | 4608.483 | 4357.293 | 4456.667 | 4344.191 |
| BIC | 4621.028 | 4476.419 | 4494.301 | 4488.395 |
| DIC(-2Log-likelihood) | 4604.484 | 4319.292 | 4444.668 | 4358.190 |

SE Standard Error; ICC: Intra-class Correlation Coefficient; MOR: Median Odds Ratio; PCV: Proportional Change in Variance; AIC: Akaike's Information Criterion; BIC: Bayesian information Criteria; DIC: Deviance Information Criterion
<sup>a</sup>Null model is an empty model, a baseline model without any explanatory variable
<sup>b</sup>Model 1 is adjusted for individual-level factors
<sup>c</sup>Model 2 is adjusted for community-level factors
<sup>d</sup>Model 3 is the final model adjusted for both individual and community-level factors
<sup>e</sup>Increased risk (in median) that one would have if moving to a neighborhood/cluster with a higher risk
\*\*\*P-value < 0.001

### Factors associated with unsafe child stool disposal

The calculated value intra-cluster correlation (ICC) was 39.61%, which indicated that the assumption of independent observation was violated (**Table 5**). Thus, we used a multilevel logistic regression model to account for the cluster effect. In the multivariable multilevel binary logistic regression model, the age of the child, wealth index, type of toilet facility, region, and community poverty level were identified as associated factors with unsafe child stool disposal. The age of the child was an important predictor of unsafe child stool disposal. Children in the age group of 6-11 months, 12-17 months, and 18-23 months were about 34% (AOR: 0.66, 95%CI: 0.52-0.83), 32% (AOR: 0.68, 95%CI: 0.54-0.86), and 42% (AOR:0.58, 95%CI: 0.45-0.74) less likely to have unsafe child stool disposal than children younger than 0-5 months, respectively. Wealth index also showed a strong statistical association with unsafe child stool. Children belonging to the poorest wealth quintiles had a four times higher chance of unsafe child stool disposal (AOR: 4.62, 95%CI: 2.98-7.16) than the richest children. Similarly, children belonging to the poorer (AOR = 2.77, 95%CI: 1.82-4.23), middle AOR: 2.13, 95%CI: 1.41-3.22), and richer wealth quintiles (AOR: 1.56, 95%CI: 1.05-2.32) had higher odds of unsafe child stool disposal than the richest children. Children belong to households who had unimproved toilet facilities were about 54% (AOR: 1.54, 95%CI: 1.17-2.02) more likely to had unsafe child stool disposal than children in households with improved toilet facilities. The likelihood of unsafe child stool disposal among children belong to agrarian regions was about 38% (AOR: 0.62, 95%CI 0.42-0.91) lower compared with city dwellers. Unsafe child stool disposal is more prevalent among households that are high community poorer level (AOR: 1.74, 95%CI: 1.23-2.46) than those with younger children live in low community poverty level (**Table 6**).

**Table 6:** Multivariable multilevel logistic regression analysis to determine associated factors of unsafe child stool disposal in Ethiopia, EDHS 2016

| Background characteristics | Null model<br>Empty<br>model | Model 1<br>Individual-level<br>variables<br>AOR (95% CI) | Model 2<br>Community-level<br>variables<br>AOR(95% CI) | Model 3<br>Individual- and<br>community-level<br>variables<br>AOR(95% CI) |
| --- | --- | --- | --- | --- |
| **Individual-level factors** |  |  |  |  |
| **Sex of the child (n=4144)** |  |  |  |  |
| Male |  | 1 |  | 1 |
| Female |  | 0.86(0.73-1.01) |  | 0.86(0.73-1.02) |
| **Age of the child** |  |  |  |  |
| 0-5 months |  | 1 |  | 1 |
| 6-11 months |  | 0.66(0.52-0.83)*** |  | 0.65(0.52-0.83)*** |
| 12-17 months |  | 0.68(0.54-0.86)** |  | 0.68(0.54-0.86)** |
| 18-23 months |  | 0.58(0.45-0.74)*** |  | 0.58(0.45-0.75)*** |
| **Diarrhea in the last two weeks (n=4129)** |  |  |  |  |
| Yes |  | 0.79(0.63-1.01) |  | 0.82(0.65-1.03) |
| No |  | 1 |  | 1 |
| **Mother educational level** |  |  |  |  |
| No formal education |  | 1 |  | 1 |
| Primary |  | 0.81(0.66-1.01) |  | 0.85(0.68-1.05) |
| Secondary |  | 1.16(0.83-1.62) |  | 1.23(0.87-1.73) |
| Higher |  | 0.97(0.62-1.52) |  | 0.99(0.63-1.56) |
| **Mother's age (4143)** |  |  |  |  |
| 15-24 |  | 1 |  | 1 |
| 25-34 |  | 0.86(0.70-1.05) |  | 0.88(0.72-1.07) |
| >34 |  | 0.82 (0.63-1.06) |  | 0.85(0.65-1.10) |
| **Mother's working status** |  |  |  |  |
| Not working |  | 1 |  | 1 |
| Working |  | 0.92(0.77-1.10) |  | 0.95(0.79-1.14) |
| **Household wealth index** |  |  |  |  |
| Poorest |  | 6.35(4.49-8.99)*** |  | 4.62(2.98-7.16)*** |
| Poorer |  | 3.28(2.32-4.63)*** |  | 2.77(1.82-4.23)*** |
| Middle |  | 2.30(1.63-3.24)*** |  | 2.13(1.41-3.22)*** |
| Richer |  | 1.60(1.15-2.24)** |  | 1.56(1.05-2.32)** |
| Richest |  | 1 |  | 1 |
| **Toilet facility** |  |  |  |  |
| Improved* |  | 1 |  | 1 |
| Unimproved |  | 1.41(1.08-1.83)** |  | 1.54(1.17-2.02)** |
| **Source of drinking water** |  |  |  |  |
| Improved** |  | 1 |  | 1 |
| Unimproved |  | 1.17(0.95-1.44) |  | 1.14(0.92-1.41) |
| **Community level factors** |  |  |  |  |
| **Region** |  |  |  |  |
| Agrarian |  |  | 0.57(0.52-1.12) | 0.62(0.42-0.91)** |
| Pastoralist |  |  | 1.34(0.84-2.13) | 0.87(0.54-1.40) |
| City |  |  | 1 | 1 |
| **Place of residence** |  |  |  |  |
| Urban |  |  | 1 | 1 |
| Rural |  |  | 2.17(1.49-3.16)*** | 1.08(0.68-1.72) |
| **Community poverty level** |  |  |  |  |
| High |  |  | 3.22(2.33-4.43)*** | 1.74(1.23-2.46)** |
| Low |  |  | 1 | 1 |

\*\*\*p-value < 0.001; \*\*p-value < 0.05; AOR: Adjusted Odds Ratio

## Discussion

This study aimed to explore the spatial variations in unsafe child stool disposal in Ethiopia. It also aimed to identify the individual- and community-level factors associated with unsafe child stool disposal practice among women using the demographic health survey (DHS) conducting in Ethiopia. Unsafe child stool disposal was found to be a spatial problem in Ethiopia. It was also found that unsafe child stool disposal was significantly associated with different individual and community level factors. At the individual level, the age of the child, wealth index, and types of toilet facilities were associated with unsafe stool disposal. At the community level, region and community poverty were the identified factors associated with unsafe child stool disposal.

In Ethiopia, the proportion of unsafe child stool disposal was 63.10%; the highest proportion was reported in rural areas (p< 0.001). This is almost consistent with a previous similar study, which found unsafe child feces disposal reported by 67% of households in Ethiopia [14]. However, this figure was lower than unsafe child faces disposal reported from India 79% [3], 72.4% [21], and Bangladesh 80% [20,25], 84% [26]. This discrepancy may due to the operational definition used in classifying unsafe child stool disposal; as the study from Bangladesh includes buried as safe child feces disposal [20]. The other possible reasons for this disparity may be due to study participants; previous studies include under-5 children and we included the youngest child under age two. On the other hand, a much lower prevalence of unsafe child stool disposal was reported from Malawi, 14.5% [15]. This could be attributed to continued efforts in raising awareness, and ODF campaigns programs in Malawi [15].

In the global spatial autocorrelation analysis of this study, a clustering pattern of unsafe child stool disposal across the study areas was observed (Global Moran’s I = 0.211, p-value < 0.0001). This indicates that unsafe child stool disposal in Ethiopia was aggregated in specific areas. Accordingly, the hot-spot areas were found in Tigray (Central, and northeast), Afar (West, and South), Amhara (Central, North, and Southeast), Gambela (West), Oromia (East), North and some parts of Somali regions. The possible explanation for geographic variation in the prevalence of unsafe child stool disposal might be due to high open defecation practice in these identified hot spot areas. According to EDHS-2016 report, one in three households in Ethiopia have no toilet facility (39% in rural areas and 7% in urban areas) and open defecation was practices in 32.9% of the households (37.7% in rural areas and 6.8% in urban areas). Closer looks in these hot spot areas showed that unsafe child stool disposal is relatively aggregated in rural areas. Consistent with this affirmation, unsafe child feces disposal is more prevalent among households that defecate in the open and those in rural areas; over three fourth of the rural households in Ethiopia (81.2%) had unsafe child feces disposal while that is true only for (45.8%) of the urban households [12]. In the community-level factors (model 2), our finding also suggested that the odds of unsafe child stool disposal were two times higher among children residing in rural areas. Additionally, the high proportion of unsafe child stool disposal in this area might be due to disparity in access to improved sanitation facilities.

Consistent with previous studies in Ethiopia [12], Malawi [15], and in rural Bangladesh [20,27], women with older children were less likely to have unsafe child stool dispose of compared with those with younger children. This could be explained by the widespread belief and perception that the stool of young children is considered harmless in many communities, including Ethiopia [2,17]. Also, this strong association can be satisfactorily explained by the fact that a shift in safe disposal practices is usually seen as children grow: children being more likely to use potties and a toilet themselves as they get older and their behaviors tend to change as they get older [2,15,28].

In this study, the wealth index was a strong predictor of statistical association with unsafe child stool. Children belonging to the poorest and poorer wealth quintiles had a higher chance of unsafe child stool disposal than the children in households with the richest wealth quintiles. This finding was consistent with other related studies [12–15,19]. This association can be easily explained by the fact that households with a high wealth index are more likely aware of the negative effects of unsafe child stool disposal, and have access to improved sanitation, and therefore practice safe child stool disposal [15]. In connection, there is also evidence in the current study; unsafe child feces disposal is more prevalent among households with high community poverty levels.

In this study, children belong to households who had unimproved toilet facilities had a statistically significant association with high odds of unsafe child stool disposal, which is in line with the findings of some previous studies [12,14,29]. Yet reports also showed that among households with improved toilets or latrines, some unsafe child feces disposal behavior was reported [22].

At the community level, children belong to agrarian regions (like SNNP and Beneshangul Gumuz regions) were less likely to have unsafe child stool disposal than city dwellers. This finding highlighted the need for strong sanitation programs to strengthen their efforts in the city administration in Ethiopia. So far, the largest Community-Led Total Sanitation and Hygiene (CLTSH) campaign and other related efforts to end open defecation have mainly targeted rural communities in Ethiopia, with only a limited focus on the management of child stool among city dwellers [30,31]. As a result, child feces management should be promoted among city dwellers in the country.

## Conclusion

This study showed that unsafe child stool disposal had spatial variability across survey clusters and regions; it was higher in the northern part of the country. Both the individual-level characteristics (child’s age, wealth index, types of toilet facility) and community-level characteristics (region and community poverty) were statistically significant predictors of unsafe child stool disposal. Hence, the health authorities could tailor effective child stool management programs to mitigate the inequalities identified in this study. It is also better to consider child stool management intervention in existing sanitation activities.

## Limitations

Despite we used nationally representative data that can enhance the generalisability of the findings and a multilevel logistic regression model, a model that accounts for the correlated nature of EDHS data. The present study has several limitations. First, due to the secondary nature of the data, the present study was limited by unmeasured confounders such as mother knowledge towards child stool disposal and other community-level factors such as social and cultural norms towards child feces management. Second, self-reported practices can be subject to bias that might underestimate true levels by underreporting socially undesirable behaviors. Additionally, in EDHS self-reported child stool disposal practices have not been validated with objective measurements such as spot check observations. Third, the cross-sectional nature of the survey does not allow the cause-and-effect relationship between independent variables and unsafe child stool disposal.

## Data Availability

The data we used which is the 2016 Ethiopian Demographic and Health Survey were obtained from the DHS program (www.dhsprogram.com) but the Dataset Terms of Use do not permit us to distribute this data as per data access instructions (http://dhsprogram.com/data/Access-Instructions.cfm). To get access to the dataset you must first be a registered user of the website (www.dhsprogram.com) and download the 2016 Ethiopian Demographic and Health Survey. 

## Data availability statement

The data we used which is the ‘2016 Ethiopian Demographic and Health Survey’ were obtained from the DHS program (www.dhsprogram.com)but the ‘Dataset Terms of Use’ do not permit us to distribute this data as per data access instructions (http://dhsprogram.com/data/Access-Instructions.cfm). To get access to the dataset you must first be a registered user of the website (www.dhsprogram.com) and download the 2016 Ethiopian Demographic and Health Survey.

## Funding

No fund was received for the present study.

## Conflicts of interest

The authors declare that there are no competing interests.

## Ethics approval

DHS Programme granted permission to download and use the data for this study after being registered and submitting a request with briefly stated objectives of the study. The Institution Review Board approved procedures for DHS public-use data sets do not in any way allow respondents, households, or sample communities to be identified. There are no names of individuals or household addresses in the data files. The geographic identifiers only go down to the regional level (where regions are typically very large geographical areas encompassing several states/provinces). Each EA (primary sampling unit) has a number in the data file, but their numbers do not have any labels to indicate their names or locations. The detail of the ethical issues has been published in the 2016 EDHS final report, which can be accessed at: http://www.dhsprogram.com/publications [10].

## Supporting information

**S1_Figure 1:** The global spatial autocorrelation based on feature locations and attribute values of unsafe child stool disposal in Ethiopia, EDHS 2016

## Notes

### Competing Interest Statement

The authors have declared no competing interest.

### Author Declarations

DHS Programme granted permission to download and use the data for this study after being registered and submitting a request with briefly stated objectives of the study. The Institution Review Board approved procedures for DHS public-use data sets do not in any way allow respondents, households, or sample communities to be identified. There are no names of individuals or household addresses in the data files. The geographic identifiers only go down to the regional level (where regions are typically very large geographical areas encompassing several states/provinces). Each EA (primary sampling unit) has a number in the data file, but their numbers do not have any labels to indicate their names or locations. The detail of the ethical issues has been published in the 2016 EDHS final report, which can be accessed at: http://www.dhsprogram.com/publications.

